# The psychological distress of parents is associated with reduced linear growth of children: evidence from a nationwide population survey

**DOI:** 10.1101/2020.02.11.20022103

**Authors:** Kun A Susiloretni, Emily R Smith, Suparmi, Marsum, Rina Agustina, Anuraj H Shankar

## Abstract

**Background:** Stunting, an indicator of restricted linear growth, has become a primary measure of childhood undernutrition due to its persistent high prevalence globally, and importance for health and development. Although the etiology is recognized to be complex, most analyses have focused on biomedical determinants, with limited attention to social factors affecting care and nurturing in the home.

**Objectives:** To identify the association between parental distress and linear growth and stunting and to examine z score loss due to any parental distress and socioeconomic, behavioral, and physiologic risk factors on for linear growth

**Design:** Cross sectional study

**Participants:** 54,261 households from the Indonesia National Health Survey 2013. Main outcome measures. Multivariate linear, logistic, and multilevel multinomial logistic regression, using survey weights, were used to assess the relationship between parental distress, as assessed by the WHO Self Reporting Questionnaire, with height-for-age z score (HAZ), stunting, and behavioral risk factors for stunting.

**Results:** Maternal, paternal and parental distress (i.e. both maternal and paternal distress) were associated with reduced linear growth of the children by 0.091, 0.13 and 0.21 z-scores, respectively. Maternal and paternal distress increased the risk of mild stunting (HAZ < -1) by 38% and 39%, and the risk of moderate stunting (HAZ < -2) by 41% and 23%, respectively. Parental stress increased the risk of moderate and severe stunting by 67% and 41%. Any parental distress accounted for 5.6% of the modeled overall loss of HAZ-score due to interactions with socioeconomic status (low maternal occupational status, low wealth, low maternal occupation) and behavioral (poor garbage sanitation) factors, rendering it amongst the more influential factors on linear growth. The modeled overall loss of HAZ-score incorporating any parental distress was associated with socioeconomic factors (26.1%) including, in descending order, low maternal occupational status, low wealth, low maternal education, low paternal occupational status, rural residence; behavioral factors (15.3%) including open garbage disposal, unimproved water disposal, paternal smoking, not using iodized salt; physiological factors (13.5%) including low maternal height, female child, paternal height, maternal mid-upper arm circumference.

**Conclusions:** These findings highlight the complex etiology of stunting, and suggest nutritional and other biomedical interventions are insufficient, and that promotion of mental and behavioral health programs for parents are essential to achieve child growth and development, and would likely foster taller, healthier, smarter, and thriving populations.

## INTRODUCTION

Linear growth restriction during childhood has profound impacts on health throughout the human life course. Short-term effects include increased mortality, morbidity, and disability. Long-term consequences include reductions in adult size, intellectual ability, economic productivity, reproductive performance, and increased risk of metabolic and cardiovascular disease. Stunting, an indicator of restricted linear growth, has become a primary measure of childhood undernutrition due to its persistent high prevalence globally, and importance for health and development [2-5]. Multiple studies of stunting have been conducted, and repeatedly identified the risk factors of low maternal stature, low birth weight, childhood gut infections, and low socioeconomic status. In accord with birth weight, a recent study added fetal growth restriction and preterm birth as risk factors and suggested they account for as much as 32% of stunted children. Additional risk in some studies include environmental factors [6-8], maternal health status, and child health status. However, these risk factors combined still cannot account for even a majority of stunted children. The etiology of restricted linear growth remains poorly understood [4, 9, 10].

Because most studies have been limited to biomedical causes of growth restriction, the impact of household socio-emotional risk factors remain poorly explored. Factors such as parental psychological distress could diminish the quality of caregiving behaviors and enhance psychological stress for the **child**, both of which may affect growth via the hypothalamic– pituitary–adrenal (HPA) axis and other pathways [11, 12]. Moreover, stress in pregnant women and children could influence nutrient metabolism or immune function, further limiting fetal and child growth and adversely affecting health [13, 14]. Psychological distress assessment in India and Vietnam using the Self Reporting Questionnaire (SRQ20), found that high maternal mental disorder contributed to child stunting and underweight status [15]. Another study in Bangladesh and Vietnam found maternal mental disorder was related to child stunting and underweight [16]. In addition, previous research in Brazil found that BMI-for-age z-scores of children were negatively associated with maternal mental disorder scores at 5–8 years postpartum [17]. However, these studies were of either limited size or conducted predominantly in groups experiencing elevated stress levels and did not collect data on many of the other known predictors of poor linear growth. As such, their findings may have limited relevance to the general population and are subject to confounding.

To address this gap in knowledge of causes of growth restriction, and to better understand links between parental stress and child linear growth, we examined data from Indonesia where the prevalence of stunting over the last 10 years has remained between 33.6% to 37.2% and the prevalence of parental mental disorders between 6.0% an 11.6% as indicated by the Indonesia National Health Survey (INHS) from 2007, 2010, 2013 and 2016 [1, 18]. Specifically, we utilized the INHS 2013 national-level dataset with complete data on psychological distress in parents, child height, and other nutritional, behavioral, and social factors associated with stunting. We examined the association between parental distress and linear growth and stunting, and z score loss due to the interaction between any parental distress and socioeconomic, behavioral, and physiologic risk factors son for linear growth.

## Methods

### Design

We used nationally representative data from the INHS 2013. This was cross sectional household survey data, designed to be representative at the national, provincial, and district level. The sampling frame consisted of 12,000 census blocks selected using probability proportional to size from 30,000 Primary Sampling Units (PSUs). There were 294,959 households visited (98.3%) of 300,000 household targeted, from 33 provinces and 497 districts/cities. The INHS 2013 collected more than 1,000 variables using household and individual questionnaires. Data collection was in 2013 and approved by the Ethics Committee for Health Research of the National Institute of Health Research and Development (NIHRD) of Indonesia. All participants gave written consent [1]. For our study, we selected data from all households comprised of two-parent families with children age 6 to 59 months, and with data on parental distress.

### Variables

The primary outcomes of this study were linear growth and stunting prevalence. Height and length were measured to the nearest 0.1 cm [1], and age was assessed from respondent statements of date of birth confirmed with legal documents such as household ID listing and birth certificate. These were used to calculate the height-for-age z-score (HAZ) using WHO Anthro version 3.2. software [19]. We excluded children with missing HAZ (7.6%) and HAZ < - 6 or HAZ > 6 (0.04%) [20, 21]. HAZ scores were classified as mild, moderate, and severe stunting using cut off points of < -1 HAZ, <-2 HAZ, and <-3 HAZ, respectively.

Parental psychological distress was assessed using the Self Reporting Questionnaire (SRQ20) developed for adults by WHO. The SRQ20 is comprised of 20 questions related to neurotic symptoms [22] and has been validated in many developing countries, including Malaysia, The Philippines, and India [23-25]. Mothers and fathers were asked to indicate whether they had experienced each of the 20 symptoms (‘yes’=1 or ‘no’=0). We calculated maternal and paternal distress scores by summing the item scores, which ranged from 0 to 20 (0 indicating ‘no distress’ to 20 indicating ‘severe distress’), and also categorized maternal and paternal distress into a binary variable using a cutoff point of 6 [1]. A score <6 indicated ‘no distress’, and ≥6 indicated ‘distress’. We created four parental distress categories: both the mother and father with no distress, only the mother with distress, only the father with distress, and both the mother and father with distress. We also built a binary distress variable using ‘no parental distress’ and ‘any parental distress’ at least distress score = 1.

Other covariates of interest included disease, physiological, health behavior, and socioeconomic factors of the child and parents. Disease factors consisted of binary variables indicating whether or not the child suffered from diarrhea, upper respiratory infection, pneumonia, or malaria in the last month. Physiological factors were mid upper arm circumference (MUAC), body mass index (BMI), height and age of the mother, height and age of the father, and child sex. Health behavior factors consisted of used of iodized salt, drinking water source, garbage disposal, water waste disposal, stool disposal, hand washing behavior, and paternal smoking. Social factors were education, occupation of mother and father, number of household members, residence, and wealth quintile (Figure 1).

**Figure 1.**
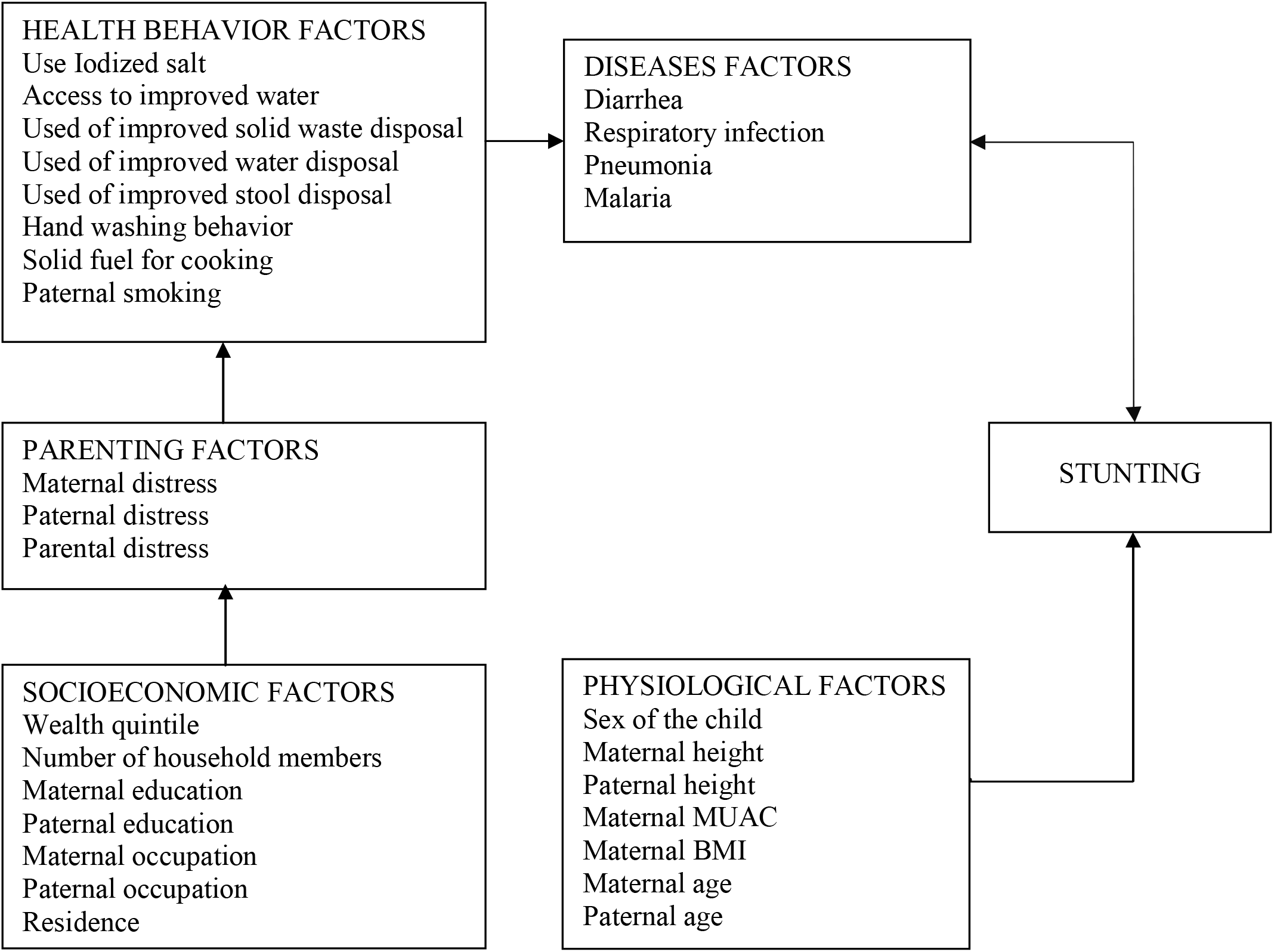
Conceptual framework of parental distress and other factors on stunting in children

### Statistical analyses

Descriptive statistics were tabulated to assess the distribution of respondents’ characteristics by each variable. The multivariate adjusted analyses incorporating sampling weights were conducted using linear, logistic, and multilevel multinomial logistic regression with PSU as a random effect. We determined the full and the best fitting model for the relative risk of stunting associated with parental distress and other covariates. We present the beta coefficient of HAZ score and proportion of z-score lost due to any parental distress and other risk factors. For the proportion of z-score lost due to any parental stress, the numerator was the sum of the products for each child of the parental risk beta coefficient times the any parental stress score, and the denominator was the sum of the z-score lost for each child based on the intercept and sum of the product of significant beta values of each covariate and the value of the response variable. We also examined the relationship between parental distress and behavioral risk factors for stunting using multilevel logistic and multinomial logistic regression. All analyses were run using STATA 13 of StataCorp. College Station, Texas.

## RESULTS

The flow diagram of study participants is shown in Figure 2. From a total of 75,440 households with children 6-59 months age, there were 54,261 households with complete parent data eligible for analysis. The mean (SD) age of the children was 34.1 (±15.4) months; fathers and mothers were 36.1 (±7.2) years and 31.6 (±6.2) years old, respectively. More than half of mothers (59.2%) were not working, while almost all fathers (96.8%) were working. We observed that 33,507 (61.8%) and 31,501 (58.1%) of the mothers and fathers had never attended a high school, respectively. The study households had a mean family size of five. Of the 54,261 children age 6-59 months, 63.0% were mildly (24.8%), moderately (21.1%), or severely (19.7%) stunted. The mean HAZ was -1.45± 2.05. The proportion of mothers, fathers, and parents with any distress (i.e. distress of at least 1) were 39.6%, 33.3%, and 51.6%, respectively, and classified as distressed (i.e. SRQ score of at least 6) were 4.1%, 2.6%, and 0.9%, respectively. Table 1 shows the distribution of variables included in the study and stratified by child stunting status (not stunted, mildly, moderately, and severely stunted).

**Table 1.** Characteristics of children and household according to stunting categories

| Factors | Normal |  | Mild stunting |  | Moderate stunting |  | Severe stunting |  | Total |  |
| --- | --- | --- | --- | --- | --- | --- | --- | --- | --- | --- |
|  | N | % | N | % | N | % | N | % | N | % |
| Stunting | 19,586 | 34.6 | 13,844 | 24.5 | 12,066 | 21.3 | 11,032 | 19.5 | 56,528 | 100.0 |
| Child Age |  |  |  |  |  |  |  |  |  |  |
| 6-11 months | 2,684 | 5.3 | 1,096 | 2.1 | 717 | 1.4 | 1,004 | 2.0 | 5,501 | 10.8 |
| 12-23 months | 4,345 | 8.5 | 2,640 | 5.2 | 2,282 | 4.5 | 2,493 | 4.9 | 11,760 | 23.1 |
| 24-35 months | 3,779 | 7.4 | 2,730 | 5.4 | 2,622 | 5.1 | 2,696 | 5.3 | 11,827 | 23.2 |
| 36-47 months | 4,116 | 8.1 | 3,466 | 6.8 | 3,009 | 5.9 | 2,584 | 5.1 | 13,175 | 25.8 |
| 48-59 months | 4,465 | 8.8 | 4,095 | 8.0 | 3,316 | 6.5 | 2,342 | 4.6 | 14,218 | 27.9 |
| Parental distress (SRQscore of at least 6) |  |  |  |  |  |  |  |  |  |  |
| No parental distress | 18,215 | 32.2 | 12,819 | 22.7 | 10,842 | 19.2 | 10,341 | 18.3 | 52,217 | 92.5 |
| Maternal distress | 654 | 1.2 | 667 | 1.2 | 581 | 1.0 | 398 | 0.7 | 2,300 | 4.1 |
| Paternal distress | 400 | 0.7 | 420 | 0.7 | 373 | 0.7 | 268 | 0.5 | 1,461 | 2.6 |
| Parental distress | 120 | 0.2 | 121 | 0.2 | 150 | 0.3 | 112 | 0.2 | 503 | 0.9 |
| Any distress (SRQscore of at least 1) |  |  |  |  |  |  |  |  |  |  |
| No distress | 10,113 | 17.9 | 6,377 | 11.3 | 5,278 | 9.3 | 5,570 | 9.9 | 27,338 | 48.4 |
| Any maternal distress | 7,002 | 12.4 | 5,924 | 10.5 | 5,189 | 9.2 | 4,235 | 7.5 | 22,350 | 39.6 |
| Any paternal distress | 5,888 | 10.4 | 4,892 | 8.7 | 4,414 | 7.8 | 3,610 | 6.4 | 18,804 | 33.3 |
| Any parental distress | 9,276 | 16.4 | 7,650 | 13.5 | 6,668 | 11.8 | 5,549 | 9.8 | 29,143 | 51.6 |
| Morbidity |  |  |  |  |  |  |  |  |  |  |
| No disease | 17,291 | 30.6 | 12,416 | 22.0 | 10,533 | 18.6 | 9,692 | 17.2 | 49,932 | 88.4 |
| 1 disease | 1,868 | 3.3 | 1,426 | 2.5 | 1,251 | 2.2 | 1,246 | 2.2 | 5,791 | 10.3 |
| 2 or more diseases | 230 | 0.4 | 185 | 0.3 | 162 | 0.3 | 181 | 0.3 | 758 | 1.3 |
| Iodized salt Used |  |  |  |  |  |  |  |  |  |  |
| Yes | 15,754 | 28.2 | 11,101 | 19.9 | 9,188 | 16.4 | 8,622 | 15.4 | 44,665 | 79.9 |
| No | 3,435 | 6.1 | 2,783 | 5.0 | 2,635 | 4.7 | 2,371 | 4.2 | 11,224 | 20.1 |
| Sex of the child |  |  |  |  |  |  |  |  |  |  |
| Girls | 9,681 | 17.1 | 7,008 | 12.4 | 5,895 | 10.4 | 5,264 | 9.3 | 27,848 | 49.3 |
| Boys | 9,708 | 17.2 | 7,019 | 12.4 | 6,051 | 10.7 | 5,855 | 10.4 | 28,633 | 50.7 |
| Maternal height |  |  |  |  |  |  | - |  | - |  |
| ≥ 150 cm | 14,741 | 26.1 | 9,537 | 16.9 | 7,019 | 12.4 | 6,865 | 12.2 | 38,162 | 67.7 |
| < 150 cm | 4,606 | 8.2 | 4,472 | 7.9 | 4,904 | 8.7 | 4,234 | 7.5 | 18,216 | 32.3 |
| Maternal BMI |  |  |  |  |  |  | - |  | - | 0.0 |
| ≥ 30 | 1,812 | 3.2 | 1,410 | 2.5 | 1,037 | 1.8 | 776 | 1.4 | 5,035 | 8.9 |
| 18.5-30 | 16,244 | 28.9 | 11,579 | 20.6 | 9,926 | 17.6 | 9,350 | 16.6 | 47,099 | 83.7 |
| ≤ 18.5 | 1,267 | 2.3 | 1,006 | 1.8 | 941 | 1.7 | 956 | 1.7 | 4,170 | 7.4 |
| MUAC |  |  |  |  |  |  |  |  |  |  |
| ≥ 23.5 cm | 16,176 | 29.2 | 11,498 | 20.7 | 9,446 | 17.0 | 8,555 | 15.4 | 45,675 | 82.4 |
| < 23.5 cm | 2,844 | 5.1 | 2,301 | 4.2 | 2,280 | 4.1 | 2,315 | 4.2 | 9,740 | 17.6 |
| Maternal age |  |  |  |  |  |  | - |  | - | 0.0 |
| <25 years | 7,362 | 13.0 | 5,330 | 9.4 | 4,550 | 8.1 | 4,380 | 7.8 | 21,622 | 38.3 |
| 25-34 years | 9,740 | 17.2 | 6,960 | 12.3 | 5,943 | 10.5 | 5,456 | 9.7 | 28,099 | 49.7 |
| ≥ 35 years | 2,287 | 4.0 | 1,737 | 3.1 | 1,453 | 2.6 | 1,283 | 2.3 | 6,760 | 12.0 |
| Paternal height |  |  |  |  |  |  | - |  | - |  |
| ≥ 155 cm | 18,027 | 32.1 | 12,843 | 22.9 | 10,579 | 18.8 | 9,640 | 17.2 | 51,089 | 90.9 |
| < 155 cm | 1,272 | 2.3 | 1,119 | 2.0 | 1,300 | 2.3 | 1,419 | 2.5 | 5,110 | 9.1 |
| Paternal age |  |  |  |  |  |  | - |  | - | 0.0 |
| <30 years | 3,453 | 6.1 | 2,378 | 4.2 | 2,076 | 3.7 | 2,142 | 3.8 | 10,049 | 17.8 |
| 30-39 years | 10,170 | 18.0 | 7,319 | 13.0 | 6,205 | 11.0 | 5,650 | 10.0 | 29,344 | 52.0 |
| ≥ 40 years | 5,766 | 10.2 | 4,330 | 7.7 | 3,665 | 6.5 | 3,327 | 5.9 | 17,088 | 30.3 |
| Used septic tank for stool |  |  |  |  |  |  | - |  | - |  |
| Yes | 11,870 | 25.0 | 8,014 | 16.9 | 6,040 | 12.7 | 5,228 | 11.0 | 31,152 | 65.7 |
| No | 5,079 | 10.7 | 3,931 | 8.3 | 3,661 | 7.7 | 3,586 | 7.6 | 16,257 | 34.3 |
|  | N | % | N | % | N | % | N | % | N | % |
| Used improved waste disposal |  |  |  |  |  |  | - |  | - |  |
| Yes | 2,126 | 3.8 | 1,218 | 2.2 | 718 | 1.3 | 701 | 1.2 | 4,763 | 8.4 |
| No | 17,263 | 30.6 | 12,809 | 22.7 | 11,228 | 19.9 | 10,418 | 18.4 | 51,718 | 91.6 |
| Used improved water disposal |  |  |  |  |  |  | - |  | - |  |
| Yes | 3,322 | 5.9 | 2,077 | 3.7 | 1,480 | 2.6 | 1,381 | 2.4 | 8,260 | 14.6 |
| No | 16,067 | 28.4 | 11,950 | 21.2 | 10,466 | 18.5 | 9,738 | 17.2 | 48,221 | 85.4 |
| Improved water |  |  |  |  |  |  |  |  |  |  |
| Yes | 19,091 | 33.8 | 13,839 | 24.5 | 11,717 | 20.7 | 10,858 | 19.2 | 55,505 | 98.3 |
| No | 298 | 0.5 | 188 | 0.3 | 229 | 0.4 | 261 | 0.5 | 976 | 1.7 |
| Washing hand behavior |  |  |  |  |  |  | - |  | - |  |
| Yes | 12,570 | 22.3 | 8,915 | 15.8 | 7,221 | 12.8 | 6,614 | 11.7 | 35,320 | 62.5 |
| No | 6,819 | 12.1 | 5,112 | 9.1 | 4,725 | 8.4 | 4,505 | 8.0 | 21,161 | 37.5 |
| Solid fuel for cooking |  |  |  |  |  |  | - |  | - |  |
| No | 14,193 | 25.1 | 9,818 | 17.4 | 7,542 | 13.4 | 6,643 | 11.8 | 38,196 | 67.6 |
| Yes | 5,196 | 9.2 | 4,209 | 7.5 | 4,404 | 7.8 | 4,476 | 7.9 | 18,285 | 32.4 |
| Paternal smoking |  |  |  |  |  |  | - |  | - |  |
| No | 7,301 | 12.9 | 4,905 | 8.7 | 3,920 | 6.9 | 3,664 | 6.5 | 19,790 | 35.0 |
| Yes | 12,088 | 21.4 | 9,122 | 16.2 | 8,026 | 14.2 | 7,455 | 13.2 | 36,691 | 65.0 |
| Wealth Quintile |  |  |  |  |  |  | - |  | - |  |
| Quintile 1 | 5,323 | 9.4 | 3,079 | 5.5 | 2,002 | 3.5 | 1,821 | 3.2 | 12,225 | 21.6 |
| Quintile 2 | 4,526 | 8.0 | 3,152 | 5.6 | 2,336 | 4.1 | 1,921 | 3.4 | 11,935 | 21.1 |
| Quintile 3 | 3,411 | 6.0 | 2,828 | 5.0 | 2,360 | 4.2 | 2,070 | 3.7 | 10,669 | 18.9 |
| Quintile 4 | 3,118 | 5.5 | 2,550 | 4.5 | 2,486 | 4.4 | 2,301 | 4.1 | 10,455 | 18.5 |
| Quintile 5 | 3,011 | 5.3 | 2,418 | 4.3 | 2,762 | 4.9 | 3,006 | 5.3 | 11,197 | 19.8 |
| Number of household member |  |  |  |  |  |  | - |  | - |  |
| ≤ 4 | 9,731 | 17.2 | 6,847 | 12.1 | 5,552 | 9.8 | 5,153 | 9.1 | 27,283 | 48.3 |
| > 4 | 9,658 | 17.1 | 7,180 | 12.7 | 6,394 | 11.3 | 5,966 | 10.6 | 29,198 | 51.7 |
| Maternal education |  |  |  |  |  |  | - |  | - | 0.0 |
| ≥ High School | 8,656 | 15.3 | 5,479 | 9.7 | 3,873 | 6.9 | 3,512 | 6.2 | 21,520 | 38.1 |
| Secondary school | 4,069 | 7.2 | 3,236 | 5.7 | 2,804 | 5.0 | 2,499 | 4.4 | 12,608 | 22.3 |
| Primary school | 4,686 | 8.3 | 3,838 | 6.8 | 3,710 | 6.6 | 3,435 | 6.1 | 15,669 | 27.7 |
| No graduation | 1,978 | 3.5 | 1,474 | 2.6 | 1,559 | 2.8 | 1,673 | 3.0 | 6,684 | 11.8 |
| Maternal occupation |  |  |  |  |  |  |  |  |  |  |
| Government Employee | 2,450 | 4.3 | 1,336 | 2.4 | 893 | 1.6 | 823 | 1.5 | 5,502 | 9.7 |
| Private sector | 1,662 | 2.9 | 1,180 | 2.1 | 951 | 1.7 | 825 | 1.5 | 4,618 | 8.2 |
| Low wages | 3,039 | 5.4 | 2,369 | 4.2 | 2,489 | 4.4 | 2,632 | 4.7 | 10,529 | 18.6 |
| Not working | 12,238 | 21.7 | 9,142 | 16.2 | 7,613 | 13.5 | 6,839 | 12.1 | 35,832 | 63.4 |
| Paternal education |  |  |  |  |  |  | - |  | - | 0.0 |
| ≥ High School | 9,339 | 16.5 | 6,020 | 10.7 | 4,366 | 7.7 | 3,927 | 7.0 | 23,652 | 41.9 |
| Secondary school | 3,815 | 6.8 | 2,928 | 5.2 | 2,498 | 4.4 | 2,309 | 4.1 | 11,550 | 20.4 |
| Primary school | 4,352 | 7.7 | 3,597 | 6.4 | 3,558 | 6.3 | 3,307 | 5.9 | 14,814 | 26.2 |
| No graduation | 1,883 | 3.3 | 1,482 | 2.6 | 1,524 | 2.7 | 1,576 | 2.8 | 6,465 | 11.4 |
| Paternal occupation |  |  |  |  |  |  | - |  | - |  |
| Government Employee | 4,970 | 8.8 | 3,040 | 5.4 | 2,090 | 3.7 | 1,765 | 3.1 | 11,865 | 21.0 |
| Private sector | 5,016 | 8.9 | 3,405 | 6.0 | 2,700 | 4.8 | 2,414 | 4.3 | 13,535 | 24.0 |
| Low wages | 7,863 | 13.9 | 6,340 | 11.2 | 6,209 | 11.0 | 6,036 | 10.7 | 26,448 | 46.8 |
| Not working | 1,540 | 2.7 | 1,242 | 2.2 | 947 | 1.7 | 904 | 1.6 | 4,633 | 8.2 |
| Residence |  |  |  |  |  |  | - |  | - |  |
| Urban | 9,752 | 17.3 | 6,550 | 11.6 | 4,928 | 8.7 | 4,160 | 7.4 | 25,390 | 45.0 |
| Rural | 9,637 | 17.1 | 7,477 | 13.2 | 7,018 | 12.4 | 6,959 | 12.3 | 31,091 | 55.0 |

Notes: MUAC: mid upper arm circumference, BMI: body mass index

**Figure 2.**
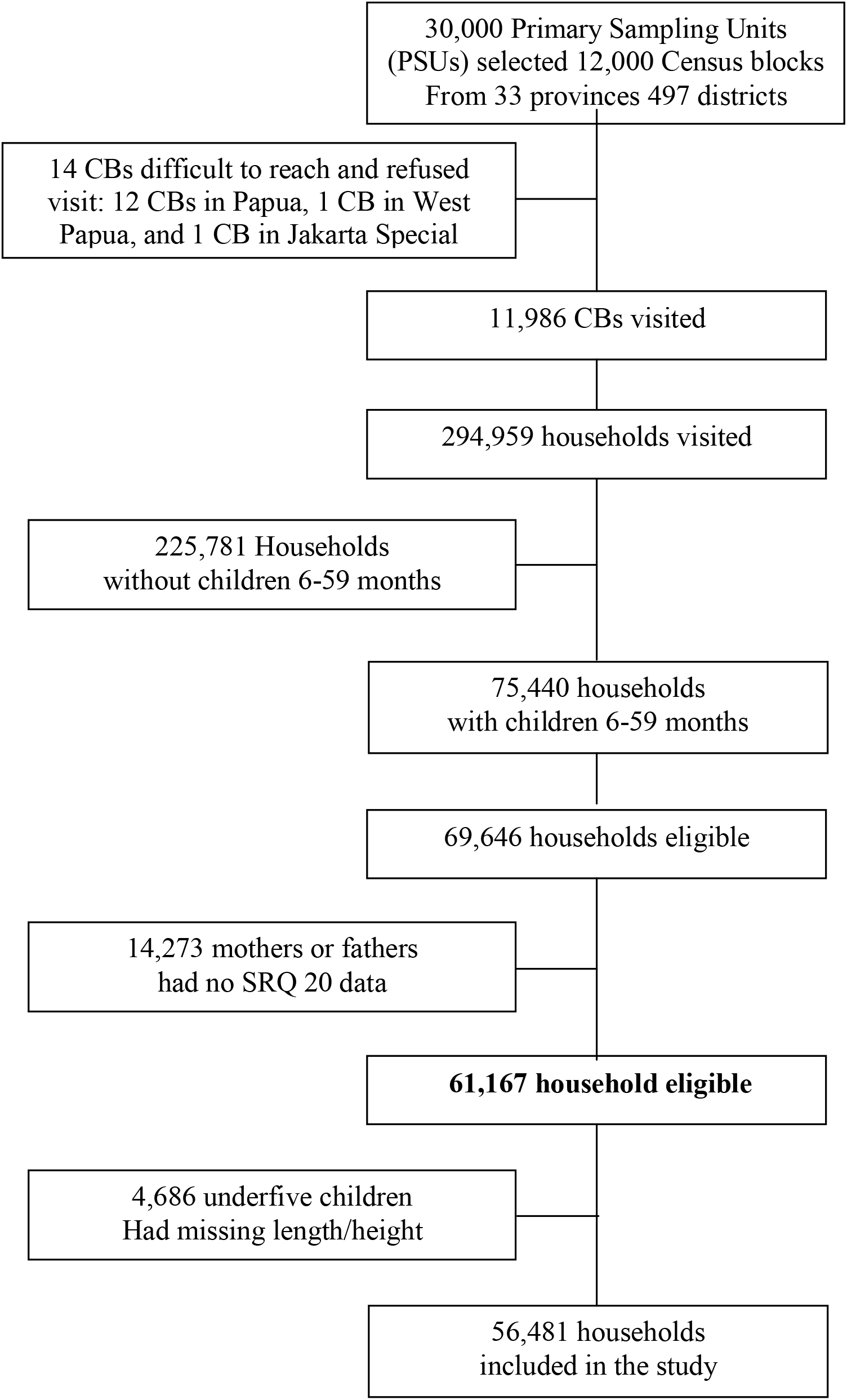
Flow diagram of participant enrollment

As seen in Figure 3, higher scores of maternal, paternal, and parental distress were associated with lower HAZ scores (p<0.05). Table 2 presents the beta coefficient of HAZ of maternal, paternal, and parental distress and other covariates grouped by social, physiological, health behavior, disease, and parental distress factors. Parental distress accounted for a reduction of 0.36 HAZ. Elevated maternal, paternal and parental distress were significantly related to lower HAZ scores by -0.09 (95%CI -0.24 to -0.023, p=0.042), -0.13 (95%CI -0.24 to -0.027, p=0.017), and -0.21 (95%CI -0.40 to -0.028, p=0.024), respectively.

**Table 2.**
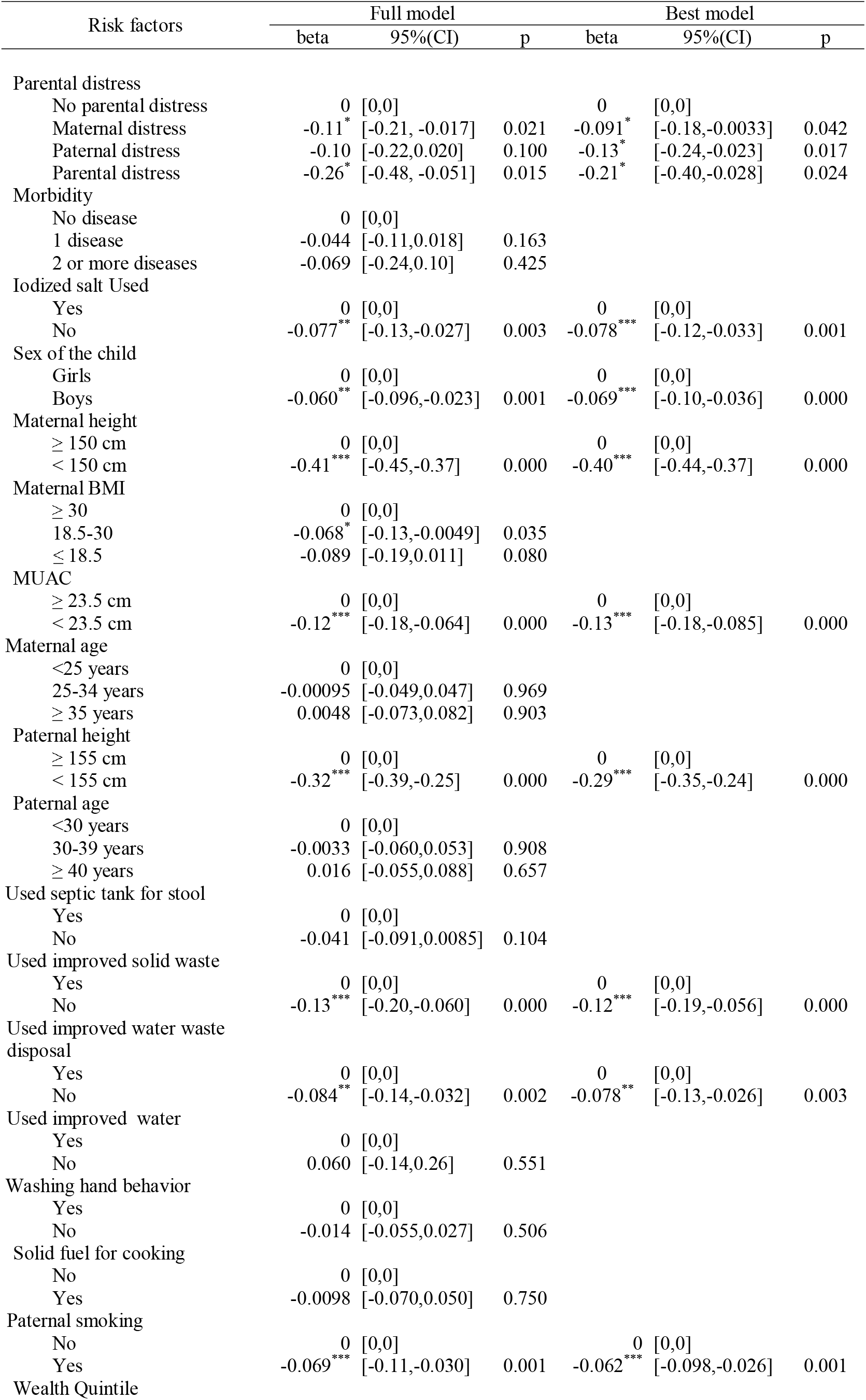
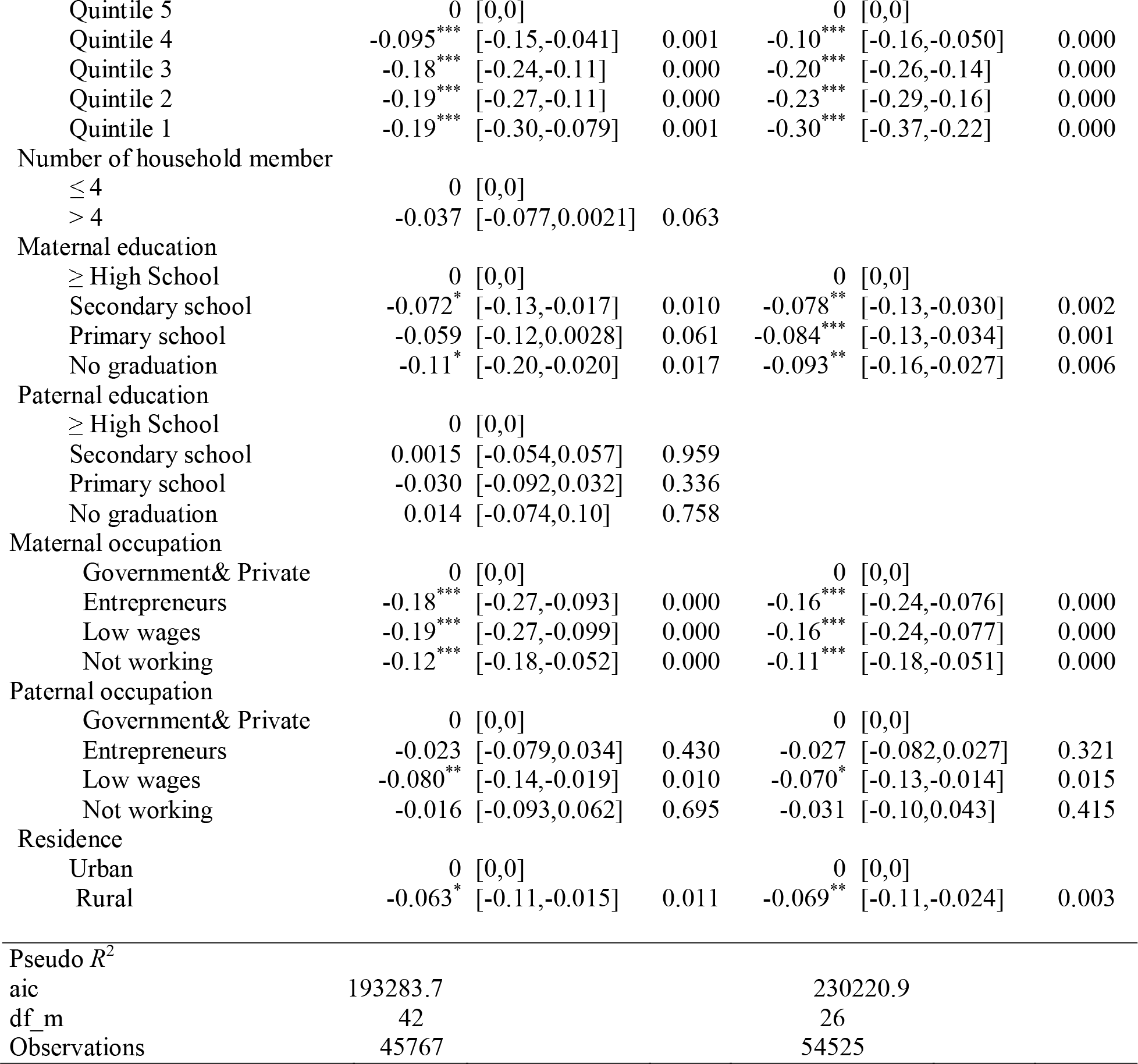
Coefficient beta of HAZ for parental distress and other risk factors in the primary sampling unit (PSU) random effect.

| Risk factors | Full model |  |  | Best model |  |  |
| --- | --- | --- | --- | --- | --- | --- |
|  | beta | 95%(CI) | p | beta | 95%(CI) | p |
| Parental distress |  |  |  |  |  |  |
| No parental distress | 0 | [0,0] |  | 0 | [0,0] |  |
| Maternal distress | -0.11* | [-0.21, -0.017] | 0.021 | -0.091* | [-0.18, -0.0033] | 0.042 |
| Paternal distress | -0.10 | [-0.22, 0.020] | 0.100 | -0.13* | [-0.24, -0.023] | 0.017 |
| Parental distress | -0.26* | [-0.48, -0.051] | 0.015 | -0.21* | [-0.40, -0.028] | 0.024 |
| Morbidity |  |  |  |  |  |  |
| No disease | 0 | [0,0] |  |  |  |  |
| 1 disease | -0.044 | [-0.11, 0.018] | 0.163 |  |  |  |
| 2 or more diseases | -0.069 | [-0.24, 0.10] | 0.425 |  |  |  |
| Iodized salt Used |  |  |  |  |  |  |
| Yes | 0 | [0,0] |  | 0 | [0,0] |  |
| No | -0.077** | [-0.13, -0.027] | 0.003 | -0.078*** | [-0.12, -0.033] | 0.001 |
| Sex of the child |  |  |  |  |  |  |
| Girls | 0 | [0,0] |  | 0 | [0,0] |  |
| Boys | -0.060** | [-0.096, -0.023] | 0.001 | -0.069*** | [-0.10, -0.036] | 0.000 |
| Maternal height |  |  |  |  |  |  |
| ≥ 150 cm | 0 | [0,0] |  | 0 | [0,0] |  |
| < 150 cm | -0.41*** | [-0.45, -0.37] | 0.000 | -0.40*** | [-0.44, -0.37] | 0.000 |
| Maternal BMI |  |  |  |  |  |  |
| ≥ 30 | 0 | [0,0] |  |  |  |  |
| 18.5-30 | -0.068* | [-0.13, -0.0049] | 0.035 |  |  |  |
| ≤ 18.5 | -0.089 | [-0.19, 0.011] | 0.080 |  |  |  |
| MUAC |  |  |  |  |  |  |
| ≥ 23.5 cm | 0 | [0,0] |  | 0 | [0,0] |  |
| < 23.5 cm | -0.12*** | [-0.18, -0.064] | 0.000 | -0.13*** | [-0.18, -0.085] | 0.000 |
| Maternal age |  |  |  |  |  |  |
| <25 years | 0 | [0,0] |  |  |  |  |
| 25-34 years | -0.00095 | [-0.049, 0.047] | 0.969 |  |  |  |
| ≥ 35 years | 0.0048 | [-0.073, 0.082] | 0.903 |  |  |  |
| Paternal height |  |  |  |  |  |  |
| ≥ 155 cm | 0 | [0,0] |  | 0 | [0,0] |  |
| < 155 cm | -0.32*** | [-0.39, -0.25] | 0.000 | -0.29*** | [-0.35, -0.24] | 0.000 |
| Paternal age |  |  |  |  |  |  |
| <30 years | 0 | [0,0] |  |  |  |  |
| 30-39 years | -0.0033 | [-0.060, 0.053] | 0.908 |  |  |  |
| ≥ 40 years | 0.016 | [-0.055, 0.088] | 0.657 |  |  |  |
| Used septic tank for stool |  |  |  |  |  |  |
| Yes | 0 | [0,0] |  |  |  |  |
| No | -0.041 | [-0.091, 0.0085] | 0.104 |  |  |  |
| Used improved solid waste |  |  |  |  |  |  |
| Yes | 0 | [0,0] |  | 0 | [0,0] |  |
| No | -0.13*** | [-0.20, -0.060] | 0.000 | -0.12*** | [-0.19, -0.056] | 0.000 |
| Used improved water waste disposal |  |  |  |  |  |  |
| Yes | 0 | [0,0] |  | 0 | [0,0] |  |
| No | -0.084** | [-0.14, -0.032] | 0.002 | -0.078** | [-0.13, -0.026] | 0.003 |
| Used improved water |  |  |  |  |  |  |
| Yes | 0 | [0,0] |  |  |  |  |
| No | 0.060 | [-0.14, 0.26] | 0.551 |  |  |  |
| Washing hand behavior |  |  |  |  |  |  |
| Yes | 0 | [0,0] |  |  |  |  |
| No | -0.014 | [-0.055, 0.027] | 0.506 |  |  |  |
| Solid fuel for cooking |  |  |  |  |  |  |
| No | 0 | [0,0] |  |  |  |  |
| Yes | -0.0098 | [-0.070, 0.050] | 0.750 |  |  |  |
| Paternal smoking |  |  |  |  |  |  |
| No | 0 | [0,0] |  | 0 | [0,0] |  |
| Yes | -0.069*** | [-0.11, -0.030] | 0.001 | -0.062*** | [-0.098, -0.026] | 0.001 |
| Wealth Quintile |  |  |  |  |  |  |

| Risk factors | Full model |  |  | Best model |  |  |
| --- | --- | --- | --- | --- | --- | --- |
|  | beta | 95%(CI) | p | beta | 95%(CI) | p |
| Quintile 5 | 0 | [0,0] |  | 0 | [0,0] |  |
| Quintile 4 | -0.095*** | [-0.15,-0.041] | 0.001 | -0.10*** | [-0.16,-0.050] | 0.000 |
| Quintile 3 | -0.18*** | [-0.24,-0.11] | 0.000 | -0.20*** | [-0.26,-0.14] | 0.000 |
| Quintile 2 | -0.19*** | [-0.27,-0.11] | 0.000 | -0.23*** | [-0.29,-0.16] | 0.000 |
| Quintile 1 | -0.19*** | [-0.30,-0.079] | 0.001 | -0.30*** | [-0.37,-0.22] | 0.000 |
| Number of household member |  |  |  |  |  |  |
| ≤ 4 | 0 | [0,0] |  |  |  |  |
| > 4 | -0.037 | [-0.077,0.0021] | 0.063 |  |  |  |
| Maternal education |  |  |  |  |  |  |
| ≥ High School | 0 | [0,0] |  | 0 | [0,0] |  |
| Secondary school | -0.072* | [-0.13,-0.017] | 0.010 | -0.078** | [-0.13,-0.030] | 0.002 |
| Primary school | -0.059 | [-0.12,0.0028] | 0.061 | -0.084*** | [-0.13,-0.034] | 0.001 |
| No graduation | -0.11* | [-0.20,-0.020] | 0.017 | -0.093** | [-0.16,-0.027] | 0.006 |
| Paternal education |  |  |  |  |  |  |
| ≥ High School | 0 | [0,0] |  |  |  |  |
| Secondary school | 0.0015 | [-0.054,0.057] | 0.959 |  |  |  |
| Primary school | -0.030 | [-0.092,0.032] | 0.336 |  |  |  |
| No graduation | 0.014 | [-0.074,0.10] | 0.758 |  |  |  |
| Maternal occupation |  |  |  |  |  |  |
| Government& Private | 0 | [0,0] |  | 0 | [0,0] |  |
| Entrepreneurs | -0.18*** | [-0.27,-0.093] | 0.000 | -0.16*** | [-0.24,-0.076] | 0.000 |
| Low wages | -0.19*** | [-0.27,-0.099] | 0.000 | -0.16*** | [-0.24,-0.077] | 0.000 |
| Not working | -0.12*** | [-0.18,-0.052] | 0.000 | -0.11*** | [-0.18,-0.051] | 0.000 |
| Paternal occupation |  |  |  |  |  |  |
| Government& Private | 0 | [0,0] |  | 0 | [0,0] |  |
| Entrepreneurs | -0.023 | [-0.079,0.034] | 0.430 | -0.027 | [-0.082,0.027] | 0.321 |
| Low wages | -0.080** | [-0.14,-0.019] | 0.010 | -0.070* | [-0.13,-0.014] | 0.015 |
| Not working | -0.016 | [-0.093,0.062] | 0.695 | -0.031 | [-0.10,0.043] | 0.415 |
| Residence |  |  |  |  |  |  |
| Urban | 0 | [0,0] |  | 0 | [0,0] |  |
| Rural | -0.063* | [-0.11,-0.015] | 0.011 | -0.069** | [-0.11,-0.024] | 0.003 |
| Pseudo R<sup>2</sup> |  |  |  |  |  |  |
| aic | 193283.7 |  |  | 230220.9 |  |  |
| df_m | 42 |  |  | 26 |  |  |
| Observations | 45767 |  |  | 54525 |  |  |

**Figure 3.**
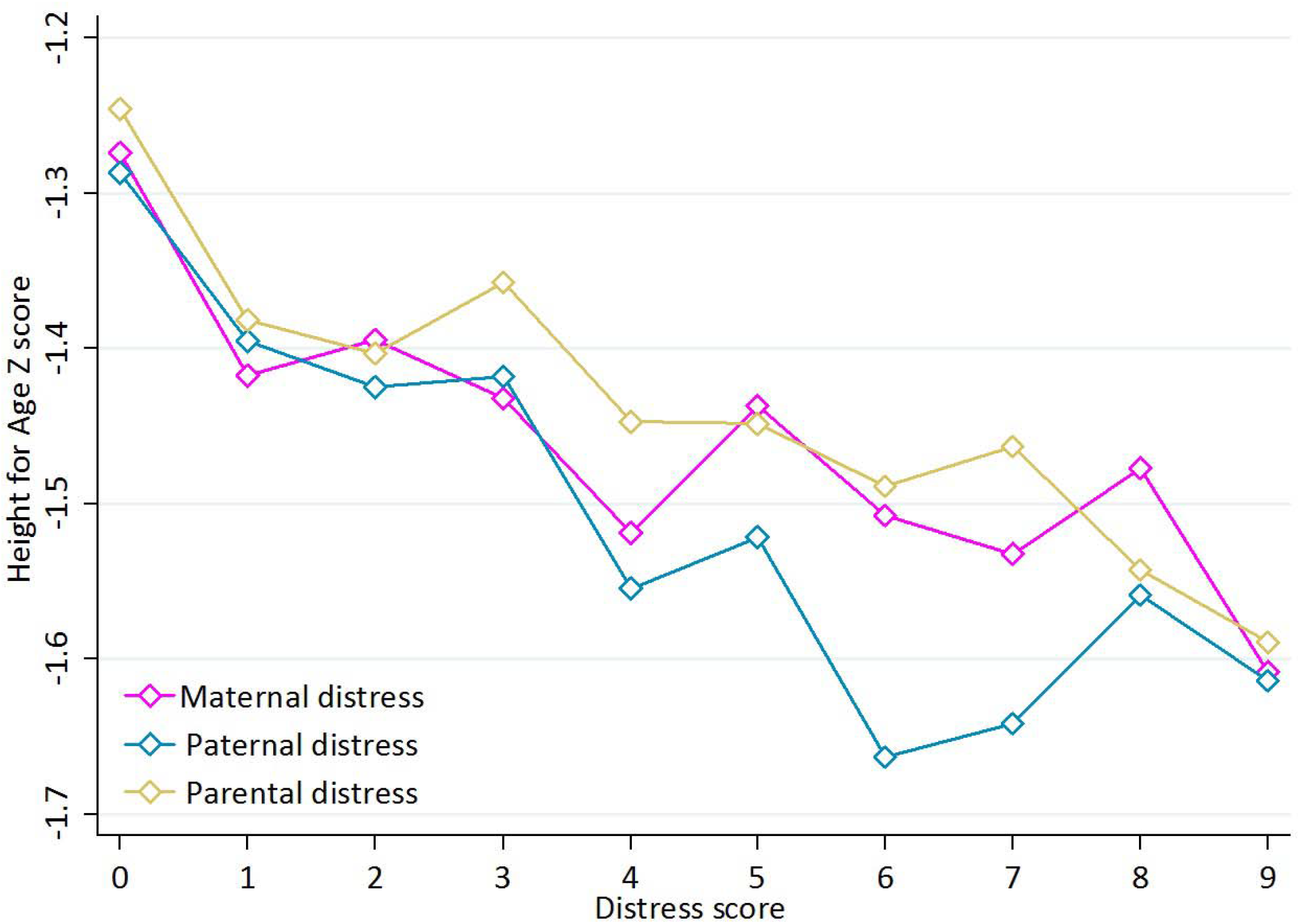
Height-for-age *Z* score by parental distress score

The relationship between parental distress and stunting is depicted in Figure 4. In the best fitting model, parental distress was a significant risk factor for mild and moderate stunting. The relative risk ratios of maternal, paternal, and parental distress for mild stunting, respectively, were 1.38 (95%CI 1.21 to 1.57, p=0.000), 1.39 (95%CI 1.18 to 1.63, p=0.000), and 1.29 (95%CI 0.94 to 1.76, p=0.114). For moderate stunting, the relative risk ratios, respectively, were 1.41 (95%CI 1.23 to 1.62, p=0.000), 1.23 (95%CI 1.03 to1.47, p=0.020), and 1.67 (95%CI 1.23 to 2.26, p=0.001). For severe stunting, the relative risk ratios for maternal, paternal, and parental stress, respectively, were 0.97 (95%CI 0.83 to 1.14, p=0.736), 1.04 (95%CI 0.86 to 1.26, p=0.671), and 1.41 (95%CI 1.02 to 1.94, p=0.036). The detailed tables and analyses are presented in Supplemental File #1.

**Figure 4.**
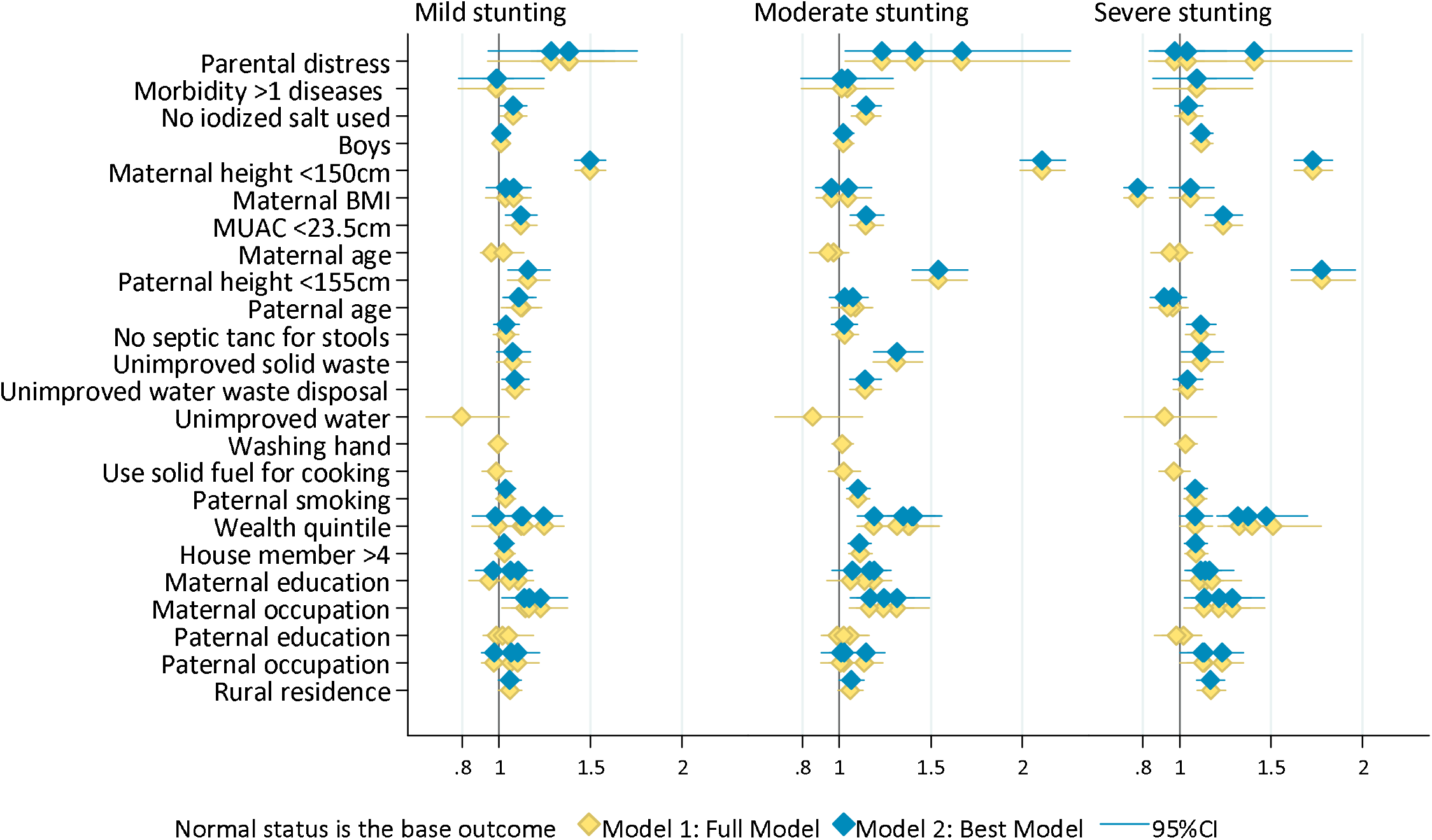
Relative risk ratio of parental distress and other covariates for stunting children age 6-59 months

Other covariates significantly related to stunting included infectious disease, physiological factors, health behaviors, and socioeconomic factors. Disease included morbidity from diarrhea, acute respiratory infection, pneumonia, and malaria. Children who experienced and least one disease were at higher risk to be severely stunted. We note that infectious disease was not associated with mild and moderate stunting.

Physiological factors related to stunting were child sex (boy), low maternal MUAC, older maternal and paternal age, paternal and maternal short stature. Boys tended to have 12% higher risk of severe stunting compared to girls (RR 1.12; 95%CI 1.07 to 1.18; p=0.000). Maternal and paternal physiological factors related to stunting including maternal and paternal height, maternal MUAC, as well as maternal and paternal age. Maternal height less than 150cm was associated with higher risk of mild (RR 1.50, 95%CI 1.341 to 1.58; p= 0.000), moderate (RR 2.11, 95%CI 1.99 to 2.23; p=0.000) and severe stunting (RR1.73, 95%CI 1.63 to 1.96; p=0.0000). Paternal height less than 155cm was associated with mild (RR=1.16, 95%CI 1.05 to 1.28; p=0.000), moderate (RR=1.54, 95%CI 1.40 to 1.70; p=0.000) and severe (RR=1.78, 95%CI 1.61 to 1.98; p=0.000). Maternal MUAC was also related to risk of mild (RR=1.12, 95%CI 1.04 to 1.21; p=0.004), moderate (RR=1.15, 95%CI 1.06 to 1.24; p=0.001) and severe (RR=1.24, 95%CI 1.14 to 1.34; p=0.000) stunting. Children of mothers with BMI≥30 had 23% lower risk of severe stunting (RR=0.77, 95%CI 0.70 to 0.85; p=0.000). Children with fathers aged ≥40 years had higher risk for mild stunting (RR=1.11, 95%CI 1.02 to 1.20; p=0.007), as did fathers age 30-39 years (RR=1.11, 95%CI 1.02 to 1.16; p=0.010), as compared to fathers aged < 30 years, but not for moderate and severe stunting.

Several behavioral factors related to stunting included non-use of iodized salt, open garbage disposal, unimproved water disposal, not having a septic tank for stools, and paternal smoking. However, households of children who had unimproved solid waste disposal, poor hand-washing behavior and use of solid fuel for cooking were not associated with stunting. Households with non-use of iodized salt had higher risk for mild (RR 1.08; CI 95%1.01 to 1.15; p=0.027) and moderately (RR 1.15, CI 95%1.07to 1.23, p=0.000) stunted children. Households who had open garbage disposal had higher risk for moderate (RR=1.32, 95%CI 1.19 to 1.46; p= 0.000) and severe (RR=1.12, 95%CI 1.01 to 1.24; p= 0.031) stunting. Households having unimproved water disposal had higher risk of mild (RR=1.09, 95%CI 1.02 to 1.16; p=0.015) and moderately (RR=1.14, 95%CI 1.06 to 1.23; p= 0.001) stunted children, but not severely stunted. Households with no septic tank for stools had higher risk for children with severe (RR=1.12, 95%CI 1.04 to 1.20; p=0.003), but not mild and moderate stunting. Children who had fathers with smoking behavior had higher risk for moderate (RR=1.10, 95%CI 1.04 to 1.17; p= 0.001) and severe (RR=1.09, 95%CI 1.03 to 1.15; p=0.005) stunting.

In addition, socioeconomic factors related to stunting were household wealth quintile, number of household members, maternal education, maternal and paternal occupation, and type of residence. The analysis showed level of wealth quintile negatively associated with mild, moderate and severe stunting. Children from the poorest quintile had 1.47-fold higher risk of severe stunting (95%CI 1.28 to 1.70; p= 0.000). The risk gradually reduced with progression to quintile 2 (RR=1.37, 95%CI 1.23 to 1.53; p=0.000), quintile 3 (RR=1.32, 95%CI 1.21 to 1.44; p=0.000) and quintile 4 (1.09, 95%CI 1.00 to 1.18; p=0.051). Lower maternal education levels, not completing high school compared to completing high school, were associated with higher risk of mild (RR=1.10, 95%CI 1.03 to 1.18; p=0.000), moderate (RR=1.19, 95%CI 1.11 to 1.28; p=0.000) and severe (1.14, 95%CI 1.06 to 1.23; p=0.001) stunting. Maternal and paternal occupation were significantly associated with stunting. Compared with children of government and privately employed mothers or fathers, children whose mothers or fathers worked as entrepreneurs, low wage employees, or were unemployed had more mild, moderate, and severe stunted children. As a private sector employee, mothers had 1.23, 1.32, and 1.22 times higher risk to have mild, moderate, and severely stunted children. Children living in rural areas had 1.06, 1.06, and 1.17 times higher risk of mild stunting compare to children living in urban areas.

Parental distress was associated with several risk factors for child stunting, including infectious disease episodes, non-use used of iodized salt, use of solid fuel, paternal smoking, use of open garbage and unimproved water disposal, and poor hand-washing behavior (Table 3). The adjusted relative risk ratios of maternal, paternal and parental distress for children who experienced one disease were 1.91 (95%CI 1.69 to 2.17, p=000), 1.43 (95%CI 1.21 to 1.69, p=001), and 2.40 (95%CI 1.87 to 2.94, p=000). Children of families with higher maternal, paternal and parental distress were at higher risk for not using iodized salt with adjusted relative risk ratios of 1.29 (95%CI 1.17 to 1.42, p=000), 1.27(95%CI 1.12 to 1.44, p=000), 1.43 (95%CI 1.17 to 1.74, p=000). Furthermore, maternal, paternal and parental distress were associated with unimproved waste disposal. However, parental distress was not related to septic tank ownership, unimproved water waste disposal and poor hand washing behavior.

**Table 3:**
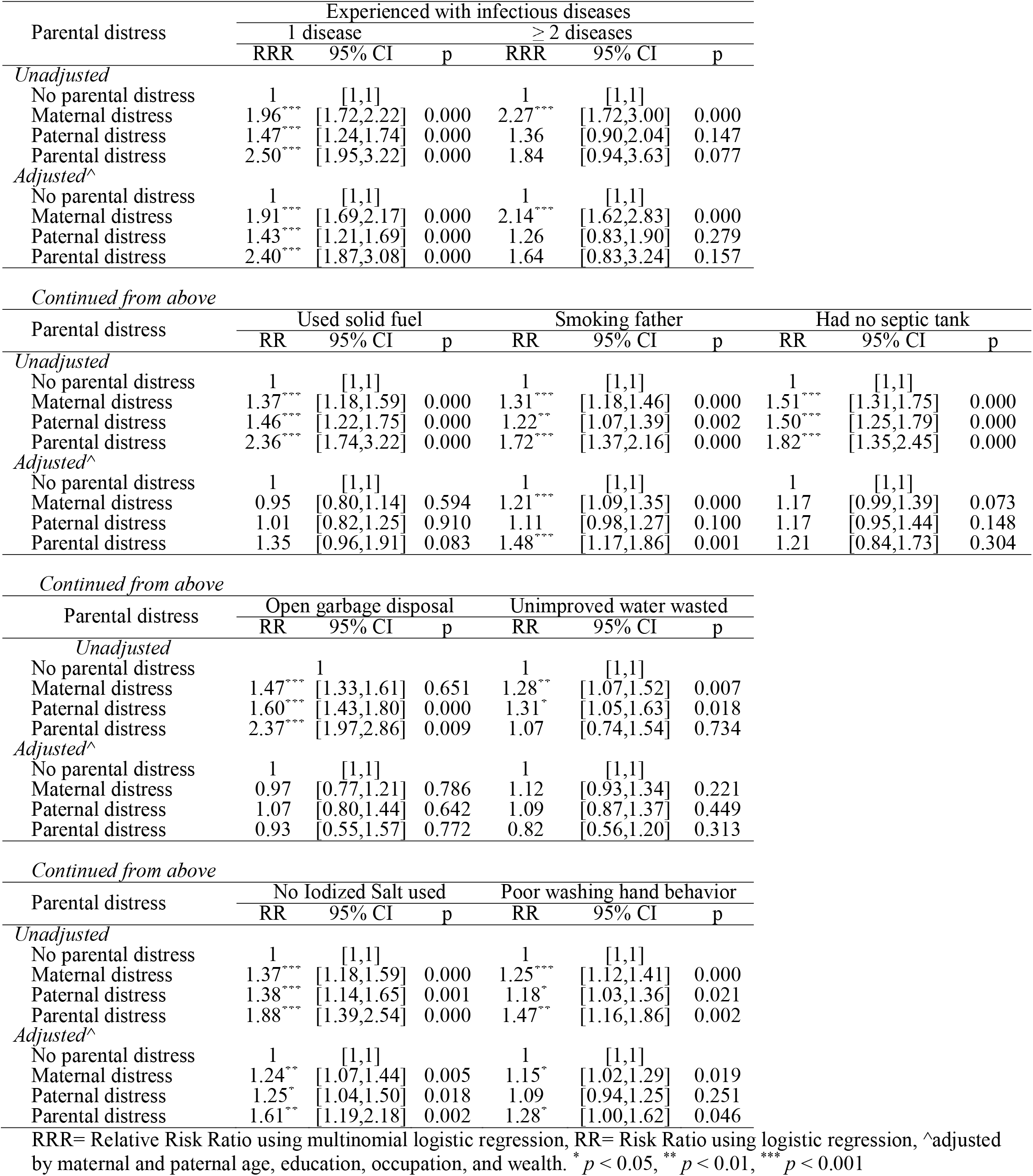
Parental distress relation to behavioral risk factors of stunting

Due to the association of distress with multiple risk factor for stunting, we used multivariate regression to model the impact of the significant predictors above, and their interaction with any parental distress, on HAZ-z score. Figure 5 and Supplemental File #2 depict the proportion of z-score lost associated with significant risk factors and their interactions with any parental distress on decreased linear growth. Amongst specific groups of risk factors, the highest proportion of z-score lost was 26.1% associated with socioeconomic factors, including low household wealth (10.5%), low maternal occupation status, low paternal occupation status, maternal education, and rural residence. The second group was behavioral factors, associated with a z-score lost of 15.3% including used of poor garbage sanitation, unimproved water disposal, smoking fathers, followed by physiological risk factors with 13.5% including maternal short stature, boys, paternal short stature, maternal MUAC, and maternal BMI.

**Figure 5.**
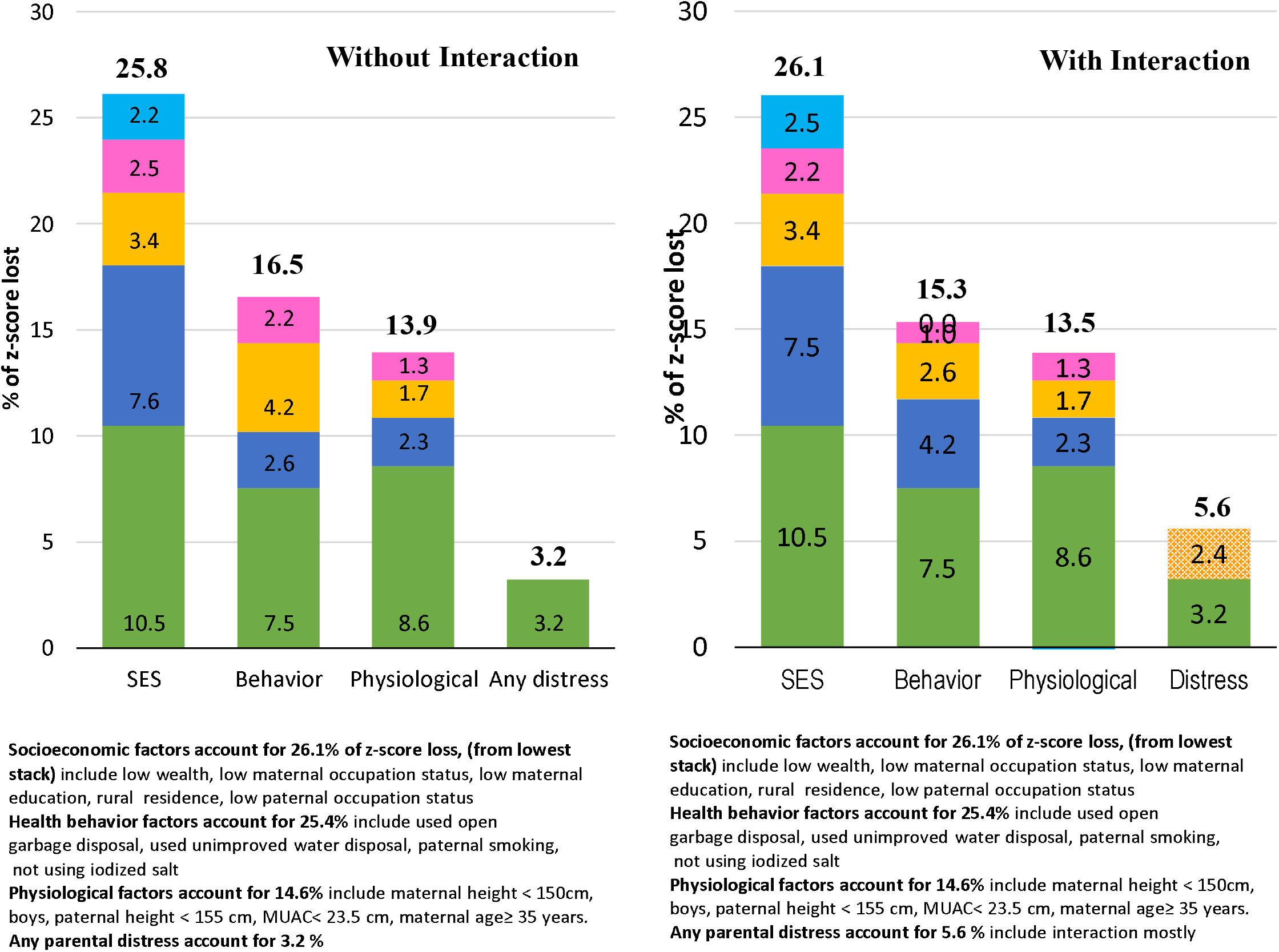
Estimate of the population proportion of z-score lost due to any parental distress and other risk factors

## DISCUSSION

We found that parental distress was associated with reduced linear growth of their children. More than half of under-five children were mildly, moderately, or severely stunted. Based on the population of Indonesia in 2013, this would account for approximately 14.65 million stunted children. And 51.6% of under-five children experienced a parent with any level of distress-either maternal (39.6%) or paternal (33.3%) or both (21.3%); and 7.5% of under-five children experienced elevated parental distress - either maternal (4.0%) or paternal (2.6%) or both (0.9%). Of those children exposed to elevated parental distress, approximately two thirds (67%) had mild, moderate or severe stunting. Based on these data we estimate approximately 11.5 million children reside in households with any parental distress, and 1.7 million stunted children in Indonesia reside in households experiencing elevated parental distress. This is a substantial burden that is likely to be underestimated. We note the proportion of households with elevated psychological distress from the Self Reporting Questionnaire (SRQ20) at the provincial level ranged from 1.6% to 15.9%, with urban centers tending toward higher stress levels, suggesting this burden is likely to increase as urbanization is increasing globally. Under-five children are in a critical stage of development, needing stimulation and attention [26, 27]. Distress among parents may affect parenting quality, especially responsiveness of their interactions, or exacerbate toxic stress that is known to affect child growth and development [28, 29].

Paternal and parental distress were associated with reductions of HAZ by 0.13, and 0.23 z-scores, respectively. This was larger than previously reported by other studies in rural Bangladesh where infants at age 12 months whose mothers has symptoms of depression had a 0.007 drop in mean HAZ [30]. A longitudinal study of a nationally representative US birth cohort showed mothers with mild and moderate depressive symptoms had children who were, on average, 0.26 cm shorter in stature (95% CI: -0.48 cm to -0.05 cm) than their peers [31].

Our analyses indicate that parental distress is related to mild and moderate stunting, but not severe stunting. The relative risk ratios of maternal, paternal, and parental distress for mild stunting were 1.23 to 1.35 and for moderate stunting were 1.25 to 1.44. This was slightly higher than reported for cohort studies in India and Peru with odds ratios of 1.18 (95% CI 1.03 to 1.35) and 1.24 (95% CI 1.07 to 1.44), respectively [32]. But the results are in line with an Indian study of maternal mental health on child stunting at 6-18 months with OR 1.4 (95%CI 1.2. to 1.6) [15], and a study from South Africa reporting an OR 1.61 (95% CI 1.02 to 2.56) [33] for stunting of children aged 2 years if their mother was socioeconomically disadvantaged and depressed (24). A higher OR of 2.17 was reported for infants of mothers with depressive symptoms in rural Bangladesh [30]. A meta-analysis found an OR was 1.4 (95%CI 1.2. to 1.7) [34]. As such, several studies support the relation of maternal mental distress or disorder on linear growth. Our analysis also examined associations of paternal distress and distress suffered by both parents on stunting and linear growth, which had not been previously reported. Paternal and parental distress showed similar association on linear growth of their children as compared to maternal distress. It is also of interest that the generation R study conducted in the high-income country of The Netherlands, which reported maternal psychological distress was positively associated with overweight children, found inconsistent associations between paternal distress and child anthropometry [35].

We found that parental distress was related to child infectious diseases and non-use of iodized salt. Parental distress may interfere with caregiving behavior which may lead to poor child growth and morbidity [36, 37]. Parents experiencing distress tend to be less engaged with their children’s daily activities and pay less attention to their growth, which also implicates less responsive feeding practice. Parents may fail to acknowledge the child’s cues of hunger, which can lead to insufficient nutrition needed to support their rapid growth and development. Also, parental distress may influence children’s stress levels, which may also influence their growth [31, 38]. Another mechanism that may increase the risk of both parental distress and stunting is poverty. Our analysis showed that socioeconomic status was linearly related to child stunting. Poverty is the source of many stunting risk factors including inadequate health care and lack of food security [36].

In our study parental distress was also associated with paternal smoking, exposure to unimproved waste and water disposal, use of unimproved water supplies, having a septic tank for stools, non-iodized salt use, and with a smoking father and poor hand-washing behavior. These would exacerbate child growth impairment. Since parenting was included in the UNICEF Framework for Improved Nutrition of Children and Women in Developing Countries in 1990 [39], there have been relatively few efforts to address this problem. This study suggests that promotion of parenting at the household level may be important for the betterment of child growth. Whereas most modifiable factors affecting undernutrition and stunting could be addressed by parenting at the household level, distressed parents might not be able to cope as well with the required actions. We recommend promotion of mental and behavioral health for parents, which would promote child growth and also promote early childhood cognitive development.

Perhaps most striking is the observation that parental distress interacting with multiple other factors accounted for 5.6% of the overall loss of HAZ-score, virtually all of its effect, rendering it amongst the larger factors in poor linear growth in this analysis. This highlights the large contribution of psychosocial factors to linear growth restriction via other factors. The other findings from this study are also important. Children suffering from even one infectious disease (diarrhea, upper respiratory tract infection, malaria) had an elevated risk for severe stunting. Physiological factors that relate to child stunting were MUAC, maternal and paternal height, maternal and paternal age, and child sex. For behavioral factors, several were found to be significant determinants of stunting. Specifically, not having a septic tank for stool disposal, using open garbage disposal, unimproved waste disposal, poor hand-washing behavior, use of unimproved water disposal, and paternal smoking were all important. The socioeconomic factors correlated with child stunting were household wealth, number of household members, maternal education, mothers’ and fathers’ occupation, and rural residence. All of those covariates have been detected by other studies [5, 40-44]. Behavior modification interventions should improve child growth by targeting improved waste water disposal, solid waste disposal, septic tank use, handwashing, and quitting smoking. While intervention on social conditions include increasing household income, improving maternal education, providing jobs, and attention to rural mothers’ conditions and working mothers.

To our knowledge, our study represents the most comprehensive analysis examining the association between parental distress from a nationally representative sample involving more than 50,000 two-parent families. Our analysis assessed maternal, paternal, and parental distress on linear growth adjusted for multiple risk factors not previously examined as group. Paternal and parental distress showed similar association with linear growth as compared to maternal distress. However, this study has limitations. First, this was a cross sectional study which could not infer causality. Second, some factors were not included in this study as predictors of linear growth including nutrition intake, child weight or length at birth, breastfeeding, food/nutrient supplementations, and feeding behavior. However, because many of these, such as birth weight and length are strongly correlated with included predictors such as maternal MUAC, maternal education, and maternal height, the findings regarding contributions of parental distress are likely to be robust.

Concerns regarding maternal distress in relation to child growth have been rising, and a better understanding is needed of interventions for maternal, paternal and parental distress. This would include more studies to understand how maternal, paternal and parental distress could reduce linear growth of the children, and therefore exacerbate the poor growth of the children. If this finding is confirmed, promotion of mental and behavioral health programs must to be pursued as part of a comprehensive strategy to enhance child growth and development.

## CONCLUSION

In summary, our analysis of nationally representative data from Indonesia found that parental distress was associated with reduced linear growth of the children and the risk of mild and moderate stunting. Moreover, parental distress was found to have associations to behavioral and other risk factors that limit child growth. As such, while the cumulative loss of height contributed from parental distress itself was limited, the effects of parental distress on overall linear growth via indirect effects is substantial. Given that multiple behavioral and socioeconomic factors influence stunting, we suggest promotion of mental and behavioral health programs for parents to promote child growth and child development.

### Research in context

#### Evidence before this study

- We identified maternal distress or depression symptoms were associated with reduced linear growth and higher odds of stunting in developing countries.
- A meta-analysis concluded that children of mothers with depression or depressive symptoms were more likely to be stunted. Studies conducted in developed countries had no consistent results.

#### Added value of this study

- To our knowledge, our study is the largest to report data about parental distress from a nationally representative sample involving more than 50,000 two-parent families. Our analysis considered both maternal, paternal, and both parent distress which was not examined in previous studies on linear growth.
- Paternal and parental distress showed similar association with linear growth compared to maternal distress. The children of parents with distresses had lower HAZ and were more likely to be stunted.
- We also compared the parental distress with other known risk factors for stunting and found that behavioral and social factors were, proportional, more important than parenting factors. Improved socioeconomic status, maternal education, and water, sanitation, and hygiene practices could avert nearly 40% of the observed stunting in the study population.
- The concern on maternal distress relation to child growth have been arise, should be followed on paternal or both parental distresses. That condition should exacerbate poor growth of the children. If this finding can be supported, a promotion of mental and behavioral health program has to be arranged to cope the concern.

## Data Availability

The technical appendix, statistical code, and dataset are available from the corresponding author.

## Footnotes

### Contributors

KAS designed the research, analyzed the data, drafted the initial manuscript, and revised the manuscript. ERS, SS, MM, and RA participated in the conception, design, data interpretation, writing and revising of the manuscript. AHS designed the research, supervised data analysis, critically reviewed and revised the manuscript. All authors have reviewed and approved the final manuscript. All authors agree to be accountable for all aspects of the work in ensuring that questions related to the accuracy or integrity of any part of the work are appropriately investigated and resolved.

### Ethical approval

Not necessary because data obtained from secondary sources that approved by the Ethics Committee for Health Research of the National Institute of Health Research and Development (NIHRD) of Indonesia.

### Data sharing

The technical appendix, statistical code, and dataset are available from the corresponding author.

